# New insights into an old vaccine: potency and breadth of neutralizing antibodies elicited by the yellow fever vaccine 17D are boosted by heterologous *Orthoflavivirus* infection

**DOI:** 10.1101/2025.07.31.25332277

**Authors:** Felicity J Coulter, Abram E Estrada, Courtney A Micheletti, Shuhua Luo, Samantha R Osman, Chad D Nix, Sophia Hu, Alisson L Naleway, William B Messer

## Abstract

The yellow fever vaccine 17D has controlled yellow fever for almost 90 years and is considered highly successful. However, with recent outbreaks in South America and Africa, yellow fever continues to represent a significant threat to public and global health. Despite its longstanding success, few studies have characterized the capacity of 17D-immune sera to neutralize wild-type (WT) yellow fever viruses (YFVs). Using a panel of WT YFVs that represent the known YFV genotypes and a unique cohort of non-endemic vaccinees with diverse *Orthoflavivirus* infection histories, we extensively characterized the potency and breadth of 17D-elicited serum neutralizing antibodies against these WT YFV. We found significant variability in neutralization test (NT) titers and rates of seropositivity of 17D-immune sera against South America-genotype I (SA-I) isolates. Vaccinee sera with serologic evidence of heterologous *Orthoflavivirus* infection had significantly great potency against SA-I strains compared to sera without evidence of *Orthoflavivirus* infection, suggesting a boosting effect from heterologous *Orthoflavivirus* immunity. These results add significantly to what is known regarding WT YFV antigenic immune landscape, including novel insights into the role of heterologous *Orthoflavivirus* infection in 17D immunity that have the potential to improve future vaccine strategies.

## Introduction

Mosquito-borne yellow fever virus (YFV) is the prototypic *Orthoflavivirus* and etiological agent of yellow fever (YF), estimated to cause up to 173,000 severe infections and up to 82,000 deaths annually across Africa and South America.^1^ With no current antiviral therapies, the live-attenuated YF vaccine 17D, developed in the 1930s, plays an essential role in controlling disease, with approximately one billion doses distributed globally^2^ which are estimated to prevent up to 119,000 cases annually.^1^

17D was derived from the WT Asibi strain, originally isolated from and named after a Ghanaian man, Asibi, with mild disease in 1927.^3^ Over the next decade, Max Theiler and colleagues serially passaged the Asibi strain first through non-human primates, and then over 180 times *in vitro* in embryonic mouse, embryonic chick, and finally embryonic chick tissues lacking brain and central nervous tissue.^4^ The resulting virus, 17D, had reduced neurotropism and viscerotropism, and did not cause disease in non-human primates; it elicited neutralizing antibodies (NAbs) and was protective against a lethal challenge with the parental Asibi strain.^4^ The first doses of 17D were administered to humans during an outbreak in Minas Gerais, Brazil in December of 1937,^5^ and the 17D vaccine rapidly became widely accepted throughout the global public health community.

Although it has a strong safety record and successfully controlled YF disease, 17D predated the modern-day FDA-approval process, bypassing contemporary phase II and III safety and efficacy clinical trials. Nonetheless, numerous studies have shown that 17D is highly immunogenic, eliciting serum NAbs in 95 to >99% of vaccinees by 30 days post vaccination^6, 7^, which are widely accepted as a correlate of protection.^8, 9^ The gold standard for characterizing 17D-elicited NAbs has been the plaque reduction neutralization test (PRNT), which determines the serum dilution at which a given percentage of input virus, for example, 50, 80, or 90%, is neutralized. Notably, most characterization of the potency and durability of 17D-immune sera have used the 17D strain^10–12^ and to a lesser degree French neurotrophic virus^15–17^ as the test virus, which both belong to the West Africa II (WA-II) genotype and have been attenuated by *in vitro* passaging.^18^ Crucially, these 17D-based PRNT data played a key role shaping recommendations^10–14^ made by the USA Advisory Committee of Immunization Practices (ACIP)^19^ despite the significant knowledge gap regarding the capacity of 17D-elicited NAbs to neutralize circulating and clinically relevant WT strains.

YFV genotypes were originally genetically defined by nucleotide sequence variation >9% within a 670 bp fragment bridging the pre-membrane (prM) and envelope (E) genes, giving rise to seven distinct genotypes.^13^ Phylogenetic analyses estimate that modern-day circulating genotypes in Africa emerged in West Africa (WA) ∼1,500 years ago, that South American (SA) strains diverged from WA strains almost 500 years, and that current circulating SA strains diverged into the SA-I and SA-II ∼300 years ago following presumed introduction into the Americas via the slave trade.^14^ The seven genotypes are WA-I, WA-II, East Africa (EA), East/Central Africa (E/CA), and Angola (A), which circulate in Africa, and SA-I and SA-II, which are prevalent in South America.

Strains belonging to the SA-I genotype caused a significant outbreak in Brazil between 2016 and 2019—the largest outbreak in the 21^st^ century^15^—with 2,205 confirmed cases and 734 deaths reported to the Pan American Health Organization^16^ although the number of actual cases was likely much higher.

To our knowledge, 17D-immune sera has been characterized using more than one authentic YFV strains only twice before: Haslwanter et al., 2022^17^ and Goncalves et al., 2024^18^ showed reduced potency of 17D-immune sera against SA-I strains amongst vaccinee cohorts in Brazil, finding that the neutralization capacity of 17D-elicited NAbs is significantly reduced against SA-I compared to 17D. Haslwanter et al. also showed reduced potency using a cohort of vaccinees from the United States (U.S.), however these data were produced using SA-I and Asibi derived reporter viral particles rather than authentic viruses. Haslwanter et al. identified five SA-I specific amino acid residue substitutions within the E glycoprotein that accounted for reduced potency of 17D-immune sera against SA-I viruses. Importantly, co-circulation of YFV, DENV, and ZIKV in Brazil and other YFV-endemic regions also elicits more complex *Orthoflavivirus* immunity within these populations that remains largely uncharacterized, and the potential for heterologous *Orthoflavivirus* infection to modify 17D-elicited NAb activity against WT YFV strains has not been explored, although this more complex immunity may have important implications for both vaccine deployment policies and future vaccine design.

To address this important knowledge gap, we assembled a panel of 13 WT YFVs including archival and contemporary strains representing all seven genotypes and immune sera from a cohort of non-endemic 17D-vaccinees with diverse *Orthoflavivirus* infection histories. Using these panels of virus and sera, we comprehensively evaluated the potency and breadth of 17D-elicited serum NAbs. Additionally, we used antigenic cartography to visualize the antigenic relationships between WT YFV strains that result from differences in susceptibility to neutralization by immune sera, providing the most extensive characterization of YFV potency and breadth to date and establishing the previously unexplored YFV antigenic landscape. By assessing geometric mean titers (GMTs) and rates of seropositivity amongst our vaccinees, we found novel potential role for heterologous *Orthoflavivirus* infection in 17D-immunity.

## Methods

### Human research ethics

This study was reviewed by and approved by the Oregon Health & Science University (OHSU) Institutional Review Board (IRB#10212 and #20910). All participants consented before enrollment.

### Study participants

All participants previously received a single dose of 17D (confirmed by vaccination record or serology) were ≥18 years of age, and recruited from the Portland metropolitan area, Oregon, USA, via two observational studies. The first cohort was recruited based on prior suspected or confirmed *Orthoflavivirus* infection or history of vaccination with 17D. The second study, conducted in collaboration with Kaiser Permanente Center for Health Research (KPCHR), recruited previously unvaccinated individuals based on upcoming scheduled 17D vaccination. Travel histories, prior infections, and information from vaccination records were documented.

### Serum

Serum was collected in serum separator tubes, allowed to clot at room temperature for ≥30 minutes before centrifugation at 1000 RCF for 10 minutes. Sera were heat inactivated at 56°C for 30 minutes and stored at -20°C or -80°C until use. *Orthoflavivirus* infection against DENV and Zika virus (ZIKV) was characterized by focus reduction neutralization test (FRNT), where an FRNT_50_ of >1:20 was considered seropositive.

### Viruses

Wildtype YFVs were obtained from the U.S. Centers for Disease Control and Prevention (CDC), the World Reference Center for Emerging Viruses and Arboviruses (WRCEVA) at the University of Texas Medical Branch (UTMB), and Betânia Drumond from the Universidade Federal de Minas Gerais. The following wildtype YFVs were used: Ogbomosho (WA-I), BA-55 (WA-I), Asibi (WA-II), Jose Cachatra (WA-II), Uganda48a (EA), BC-7914 (EA), Couma (ECA), 14FA (A), Br/MG/2001 (SA-I), BeH622205 (SA-I), 614819 (SA-I), 321/Br/MG/2018 (SA-I), and HEB4263 (SA-II). 17D was propagated from a vial of YF-VAX®.

### Virus sequencing

Viral RNA was extracted using the Quick-RNA Viral Kit (Zymo Research Corporation, catalog #R1034). Complementary DNA (cDNA) was synthesized using the primer YFVAmp2A and the SuperScript III First-Strand Synthesis System (Thermo Fisher,catalog #18080051). To synthesize cDNA, 1μL 10mM YFVAmp2A, 1μL 10mM dNTP Mix, 10μL RNA, and 13μL DEP-C treated water were combined, mixed, and incubated at 65°C for 5 minutes, then placed on ice for 2 minutes. A separate mix of 2μL 10x RT Buffer, 4μL 25mM MgCl2, 2μL 0.1M DTT, and 1μL 40 U/μL RNase OUT was prepared, added to the previous mix, and incubated at 25°C for 2 minutes. 1μL 200 U/μL SuperScript III RT was then added to each tube and incubated in the thermocycler under the following conditions: 50°C for 1 hour, then 85°C for 5 minutes. PCR amplification of the prM, E and NS1 genes was performed using Phusion High-Fidelity DNA Polymerase (New England Biolabs) and primers YFVAmp1A and YFVAmp2A. Fifty μL reactions were prepared, composed of 5μL cDNA, 2.5μL 2000 U/mL Phusion polymerase, 2.5μL 10μM forward primer (YFVAmp1A), 2.5μL 10μM reverse primer (YFVAmp2A), 1μL 10mM dNTPs, 1.5uL 100% DMSO, 10μL 5x Phusion HF Reaction Buffer, and 25μL molecular biology-grade water. Thermocycler conditions were as follows: 98°C for two minutes, 35 cycles of 98°C for 10 seconds, 60°C for 20 seconds, and 72°C for two minutes and 18 seconds, followed by 72°C for 5 minutes. PCR products were visualized on 1% agarose gel with SYBR Safe DNA gel stain (Thermo Fisher, catalog #S33102). The band of interest (approximately 3.5kb) was excised and purified using the QIAquick Gel Extraction kit (Qiagen, catalog #28704) per the manufacturer’s instructions. Purified products were sequenced commercially (Genewiz), via Sanger sequencing. Primers 857R_YFV, 996F_YFV, 1300F_YFV, 1520R_YFV, 2144F_YFV, 2340R_YFV, 2843F_YFV, 3010R_YFV, YFVAmp1A, and YFVSeq2A (Supplemental Table 1) were used to sequence the region of interest. Additional primers, 1805F_Couma, 2466R_Couma, and 3011R_Couma were designed specifically for the Couma YFV strain due to poor performance in some areas of the sequence. Contigs were assembled from .ab1 files and aligned and contigs assembled using Geneious Prime (version 2024.0.7). Geneious Prime (version 2024.0.7).

### Phylogenetic analysis

Phylogenetic analysis was conducted in MEGA11.^19, 20^ The evolutionary history was inferred from the inferred full-length E protein sequence (493 residues) using the Maximum Likelihood method and Dayhoff matrix-based model.^21^ Initial trees for the heuristic search were obtained automatically by applying Neighbor-Join and BioNJ algorithms to a matrix of pairwise distances estimated using the Dayhoff model and then selecting the topology with superior log likelihood value. A discrete Gamma distribution was used to model evolutionary rate differences among sites (5 categories (+G, parameter = 0.7260)). The tree is drawn to scale, with branch lengths measured in the number of substitutions per site.

### Virus propagation

YFVs were handled within a biosafety level 3 (BSL-3) facility at OHSU, except for 17D which was handled at BSL-2. Viruses were propagated in Vero E6 cells (ATCC, CRL-1586 ￼ transfected with a plasmid encoding human furin, although furin was found to not be overexpressed. T-75 tissue culture flasks (Fisherbrand, FB012937) were seeded with 5.0E+06 cells in 12mL of media (MEM/EBSS (Hyclone, SH30024.01), 1% v/v non-essential amino acids (NEAA; Gibco, 11140-050), 1% v/v antibiotic-antimycotic (Gibco, 15240-062), 10% v/v heat-inactivated fetal bovine serum (HI-FBS; Avantor, 89510-186), and 460µg/mL G418 (Thermo, 10131035)) and incubated at 37°C, 5% CO_2,_ overnight. Near confluent flasks were washed with 25mL of phosphate-buffered saline (PBS; HyCloneVirus was thawed at room temperature (RT) or reconstituted from lyophilized cultures with dilution media (DM; MEM/EBSS, 1% v/v NEAA, 1% v/v antibiotic-antimycotic, 2% v/v HI-FBS), added to flasks in 4mL of DM, and rocked every 15 minutes for 1 hour before the inoculum was discarded and 12mL of Vero complete media (VCM; MEM/EBSS with 10% FBS, anti-anti, L-glutamine) added. Flasks were monitored for cytopathic effects (CPE), and tissue culture supernatants were harvested when CPE was ≥40%. Tissue culture supernatants were clarified at 1000 RCF for 10 minutes at room temperature, resuspended in 10% v/v sucrose phosphate glutamate buffer (SPG; 2.18M sucrose, 38mM KH_2_PO_4_, 72mM K_2_HPO4, 60mM L-Glutamic acid), and stored at -80°C for ≥48 hours before use. Seed stocks (p1) were propagated with an unknown multiplicity of infection (MOI) using 100µL of virus suspension and used to propagate working stocks (p2) using a MOI of 0.01. Viruses were harvested 4-6 days post-infection. All assays described herein were performed using single batch p2 virus stocks.

### Focus reduction neutralization test (FRNT)

Methods were adapted from established in-house protocols used for 17D, DENV, and ZIKV.^22^ Vero cells were seeded in 96-well plates at 2.0E+04 cells per well in 200mL of VCM, and incubated at 37°C, 5% CO_2_, overnight. Using U-bottom plates, participant serum (6mL) was diluted 4-fold in DM starting at 1:5 through 1:1280 (working dilution), for a final volume of 45mL per well. Biological duplicates of serum dilutions were prepared for each virus, with a virus-only control well per each serum dilution. Viruses were thawed at RT and diluted to 2X in DM to obtain 40-150 foci per well. Virus-serum inoculums were made by adding 45mL of virus to the diluted serum (1:1) to give final serum dilutions of 1:10 through 1:2560 which were incubated for 45-60 minutes at 37°C, 5% CO_2_ to allow binding of NAbs to the surface of virions. Prior to infecting overnight cell monolayers, culture media was discarded, and 30mL of virus-serum inoculum was added to wells, followed by incubation for one hour at 37C, 5% CO2, and rocked every 15 minutes. Cells were overlaid with OptiMEM (Gibco, 31985-070) supplemented with 1% NEAA, 1% antibiotic-antimycotic, 2% v/v FBS and 5 g methylcellulose (Sigma-Aldrich, M0512-1KG). Infected plates were incubated for 48±6 hours for Couma strain and 68±4 hours for all other viruses. On day 2 or 3 post-infection, overlay media was removed, washed with PBS and fixed with 4% paraformaldehyde (Sigma-Aldrich, P6148) before removing from the BSL-3. Plates were incubated overnight at 4℃ with 150µL of blocking buffer (BB; PBS with 2% heat-inactivated normal goat serum (Rockland Immunochemicals Inc., 0204-00-0100), and 0.4% Triton-X) per well and then incubated overnight at 4℃ with 30µL of 1mg/mL 4G2 primary antibody (hybridoma-derived ATCC cat #HB-112) diluted 1:750 in BB. Plates were washed with PBS and incubated for 45-60 minutes with 30µL secondary horseradish peroxidase goat-anti-mouse-IgG antibody (BioLegend, Cat# 405306) diluted 1:1000 in BB and subsequently protected from light. Plates were washed three times with PBS and 30µL of TrueBlue™ Peroxidase Substrate (Kirkegaard & Perry Laboratories Inc., 5510-0050) added per well. Plates were developed for ≥20 minutes until foci were clearly visible by eye (occasionally overnight), quenched with water and air-dried for 20 minutes. Image and foci count data were acquired using CTL ImmunoSpot 7.0.26.0 (Cellular Technology LTD, Cleveland, OH, USA). FRNT_50_ were calculated in GraphPad Prism® (version 10.3.0) using a sigmoidal dose-response curve fitting percent neutralization against serum dilution.

### Antigenic cartography

Antigenic maps were constructed as previously described^22–24^ using FRNT_50_ data produced for our core 36 participants against 17D and 12 wildtype viruses, using the Racmacs package in R (version 4.3.1) and RStudio (version 2023.06.1+524).

### Statistical methods

Statistical analyses were conducted in GraphPad Prism® (version 10.3.0) and JMP® (version 17.2.0). After establishing that our log-transformed GMT data was not normally distributed, specifically for SA-I strains which skew-right, we employed non-parametric statistical analyses. Friedman test with Dunn’s multiple pairwise comparison was used to compare pooled GMTs Chi-squared was used to compare the proportion of seropositive and seronegative vaccinees for each virus, and Mann-Whitney U test was used to compare GMTs between 17D-only and heterologous vaccinees for each virus. For analysis of Dayhoff values against GMTs, we first established significance using a Spearman rank correlation and then applied Pearson correlation as an indicator of fit.

### Results Viruses

A total of 14 viruses were used for characterizing potency and breadth (Figure 1). The E gene of working stocks of the 14 viruses evaluated in this study were sequenced using Sanger sequencing and phylogenetic relatedness was confirmed (Figure 1A). We identified 33 amino acid differences across all viruses, with 10 differences restricted to 17D and 23 differences between WT viruses (Figure 1B). Seven differences were within E domain (ED) I, 16 were within EDII, and 10 were within EDIII (Figure 1B and C). We confirmed the presence of a novel H67N site and additional sites previously implicated by Haslwanter et al., 2022 to influence neutralization by 17D-immune sera—A83E, D270E, N271S, and N272K—which are all unique to the SA strains, with all five changes in SA-I strains apart from N271S which is also found in SA-II.

**Figure 1:**
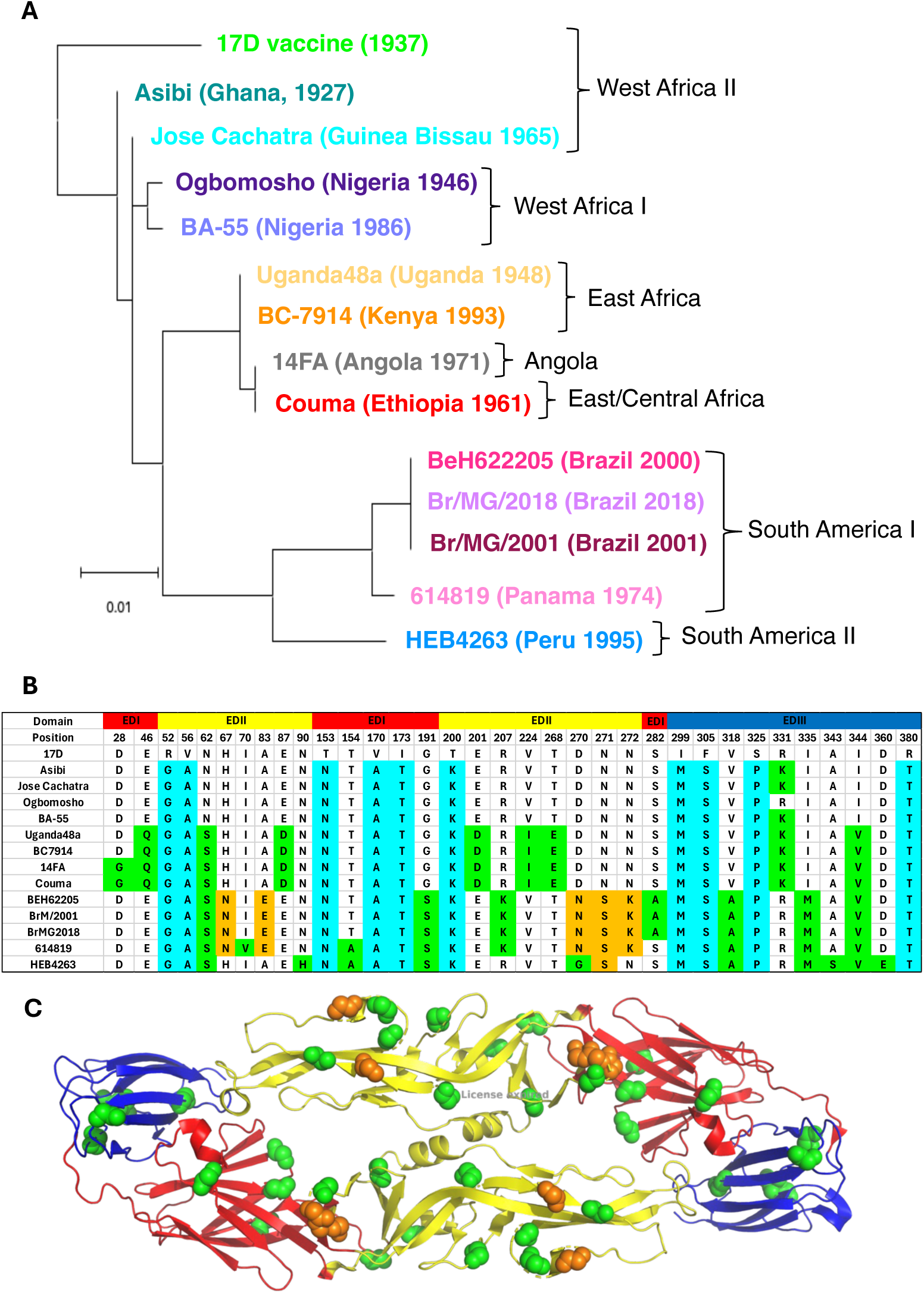
Amino acid changes within the ectodomain of the envelope glycoprotein amongst wild-type YFV strains. (**A)** Phylogenetic tree constructed using the the amino acid sequence of the E protein. Virus strain names are colored, and year of isolation is given in parentheses. Genotype names are in black. **(B)** Table of amino acid variation within the E glycoprotein ectodomain between 17D and wild-type strains, with 33 total amino acid changes, 10 restricted to 17D (cyan) and 23 between wild-type viruses (green and orange). Residues implicated in modifying neutralization (Haslwanter et al., 2022) restricted to SA-I strains are shown in orange. **(C)** Ribbon cartoon of YFV dimer with wild-type variable sites corresponding to **(B)** shown in green and orange, respectively.

WA-I strains showed the least variation with 10-11 amino acid changes (2.5-2.8%), followed by WA-II strains with 11 amino acid changes (2.8%). EA strains differed by 18-19 amino acids (4.5-4.8%), the E/CA, A, and SA-I strains each differed by 19 amino acids (4.8%), and SA-II strains showed the most variation with 22-23 amino acid changes (5.5-5.8%).

### Immune sera

The 36 study participants (Table 1) had previously received a single dose of 17D. The interval post vaccination ranged 1-11 years (median 6). Age at vaccination ranged 19-69 years (median 29.5). Vaccinees were either flavivirus naïve (n=18; “17D-only vaccinees”) or showed serological evidence of heterologous flavivirus infection (n=18; “heterologous vaccinees”).

**Table 1:** Study participants. Summary demographics of study participants, including sex, race/ethnicity, age at vaccination, years’ post-vaccination, and Orthoflavivirus infection history.

|  |  |
| --- | --- |
| Sex, n (%) |  |
| Female | 28 (77.8) |
| Male | 8 (22.2) |
| Race/Ethnicity, n (%) |  |
| White | 30 (83.3) |
| Other | 5 (13.9) |
| Unknown | 1 (2.3) |
| Age (years) at vaccination, median (min-max) | 29.5 (19-69) |
| Years post vaccination, median (min-max) | 6 (1-11) |
| *Orthoflavivirus* infection history, n (%) |  |
| 17D only vaccinees | 18 (50.0) |
| Heterologous vaccinees | 18 (50.0) |
| DENV-1 | 3 (16.7) |
| DENV-2 | 6 (33.3) |
| DENV-3 | 1 (5.6) |
| DENV-4 | 2 (11.1) |
| DENV secondary | 3 (16.7) |
| ZIKV | 2 (11.1) |
| DENV and ZIKV | 1 (5.6) |

Heterologous vaccinee immune profiles included serologic evidence of primary DENV infection to DENV-1 (n=3), DENV-2 (n=6), DENV-3 (n=1), and DENV-4 (n=2), secondary DENV (n=3), ZIKV infection (n=2), and DENV and ZIKV infection (n=1). Most participants were female (78%, n=28). Eighty-three percent of participants identified as white (n=30), 5% reported another race/ethnicity or “other” (n=5, grouped for anonymity) and 2% (n=1) is of unknown race/ethnicity.

### Potency and breadth of vaccinee immune sera

To characterize the potency and breadth of 17D-immune sera, we conducted FRNTs for each of the 14 viruses against each of the 36 immune sera. We evaluated 50% and 90% FRNT (FRNT_50_, FRNT_90_) titers and found that FRNT_50_ titers provided a more informative dynamic range as 49% of tested virus-serum combinations had FRNT_90_ titers <1:10 against WT viruses. Fold-differences for pooled GMTs of each WT virus ranged from 1.0-10.4-fold compared to 17D (Figure 2A and D). Friedman test with Dunn’s multiple comparison using 17D-serum GMTs to every other WT-serum GMT found that Uganda48a (adj. p=0.0011), 14FA (adj. p=0.0130), Couma (adj.p=0.0061), HEB4263 (adj. p<0.0001), Br/MG/2001 (adj. p<0.0001), BeH622205 (adj. p<0.0001), 614819 (adj. p<0.0001), and Br/MG/2018 (adj. p<0.0001) are significantly reduced compared to 17D while GMTs against Asibi, Jose Cachatra, Ogbomosho, and BC-7914 are not (Figure 2D). No significant differences were observed between sera-virus titers within the same genotype. Notably, SA-I strains—Br/MG/2001, BeH622205, 614819, and Br/MG/2018— FRNT_50_ titers were significantly lower compared to most other viruses, except HEB4263. (Figure 2D).

**Figure 2:**
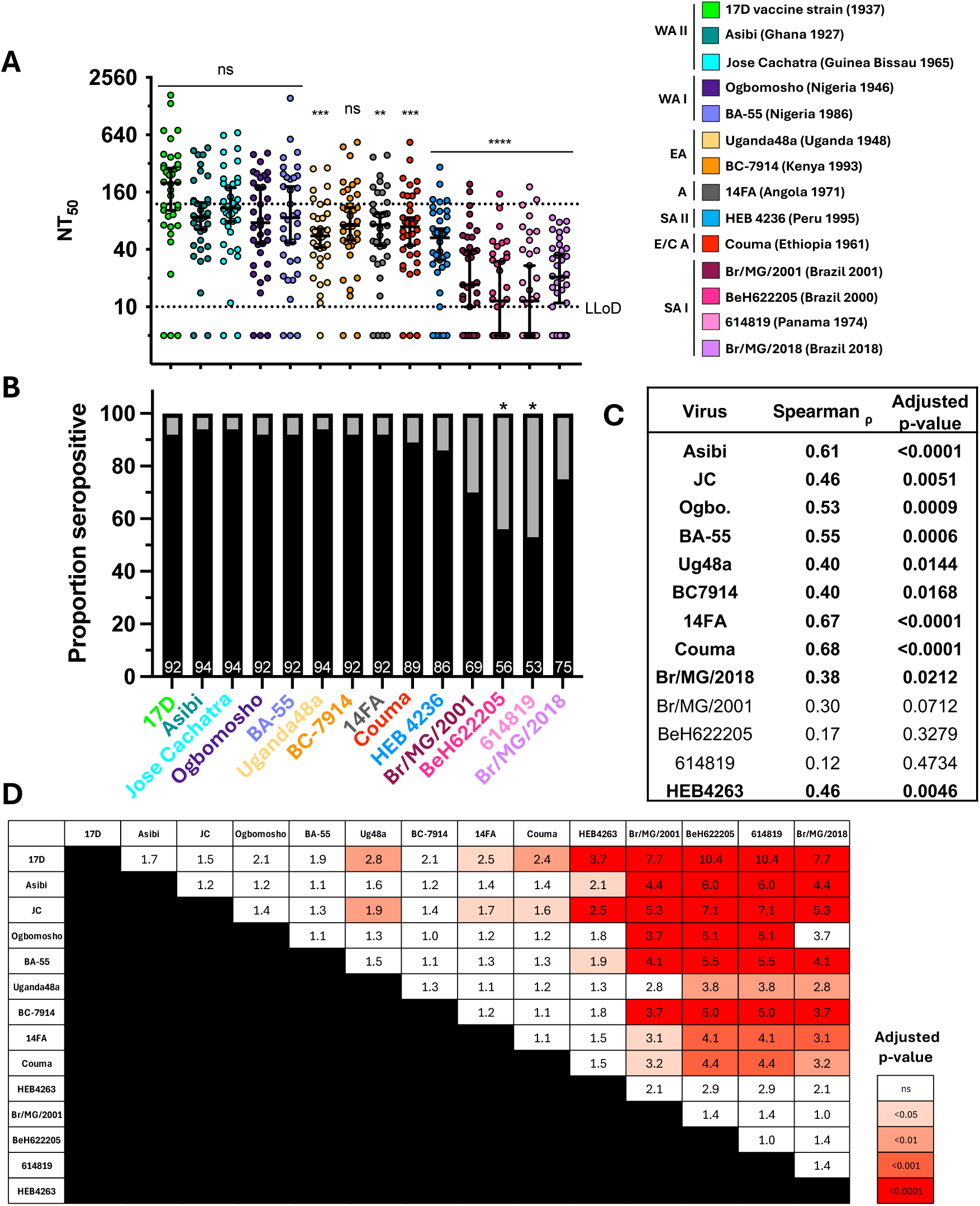
Potency of vaccinee sera against wild-type viruses are reduced compared to 17D. **(A)** NT_50_ for all participants against each virus. Each dot represents the geometric mean titer (GMT) of a single participant sera, calculated from biological duplicates. Horizontal bars and whiskers are geometric means with 95% confidence intervals. Lower limit of detection (dotted line) is 1:10 and upper limit of detection is 1:2560. GMTs <1:10 were given an arbitrary value of 5. Virus strains are color-coded according to the key and arranged by genotype. P-values derived from Friedman test with Dunn’s multiple comparison: ** adj. p<0.01, *** adj. p<0.001, **** p<0.0001. **(B)** Bar graph showing proportion of seropositive (NT_50_ ≥1:10, black) and seronegative (NT_50_ <1:10, grey) participants, with percentage of total (n=36) shown with white numbers. Chi-squared analysis shows statistically greater proportion of seronegative participants for SA-I strains Br/MG/2001, BeH622205, and 614819 compared to the model prediction. **(C)** Table showing Spearman’s rank correlation coefficients and adjusted p-values between Log-GMTs of 17D and WT viruses. Statistically significant values are in bold. **(D)** Matrix showing GMT fold differences between viruses. Boxes are colored by Friedman test with Dunn’s multiple comparisons test adjusted p-values.

We next determined proportion of vaccinees seropositive for each virus, using a cut-off of an FRNT_50_ <1:10 (Figure 2B). We found >90% of vaccinees were seropositive for all virus strains except SA-I and SA-II strains with 86% percent seropositive against HEB4263, 75% seropositive against Br/MG/2018, 69% against Br/MG/2001, and a significantly lower proportion of seropositive with 56% against BeH622205, a 53% against 614819.

Neutralization titers (NTs) against 17D are widely accepted as a correlate of protection, yet correlation of 17D-against WT YFV NTs has not been previously evaluated. To address this knowledge gap, we analyzed the correlation between 17D and WT GMTs finding that 17D FRNT_50_ titers were significantly correlated with all WT YFVs (Spearman ρ ranging 0.40-0.68, p<0.0001–0.0168) except SA-I strains, for which we found no correlation (Figure 2C).

We next examined the relationship between virus E glycoprotein primary amino acid sequences and potency of serum NAbs using Dayhoff distances. Dayhoff distances^25^ are a weighted measure of amino acid similarity between primary amino acid sequences. We found a highly significant negative correlation (Pearson’s r = -0.84, p=0.0002) between WT virus amino acid distance from 17D and WT virus neutralization titers, suggesting that as antigenic similarity to 17D decreases, serum antibody potency decreases as well.

### Heterologous vaccinees have increased potency of NAbs against SA-I strains and increased breadth of neutralization

The impact of heterotypic *Orthoflavivirus* on 17D-elicited immunity against WT YFV NAbs has not, to our knowledge, been previously examined. To explore potential effects of heterologous *Orthoflavivirus* infection history on GMTs, we stratified FRNT_50_ results by 17D-only and heterologous vaccinees (Figure 3). We found that GMTs of heterologous vaccinees were significantly higher compared to 17D-only vaccinees against all SA-I strains (p<0.0001) and individually against strains Br/MG/2001 (p=0.0109), BeH622205 (p=0.0165) and 614819 (p=0.0091) GMTs against Br/MG/2018 trended higher amongst heterologous vaccinees although this difference was not significant (p=0.2284). With no other significant differences observed, this demonstrates that heterologous vaccinees have increased potency of NAbs against SA-I strains and increased breadth of NAbs overall compared to 17D-only vaccinees.

**Figure 3:**
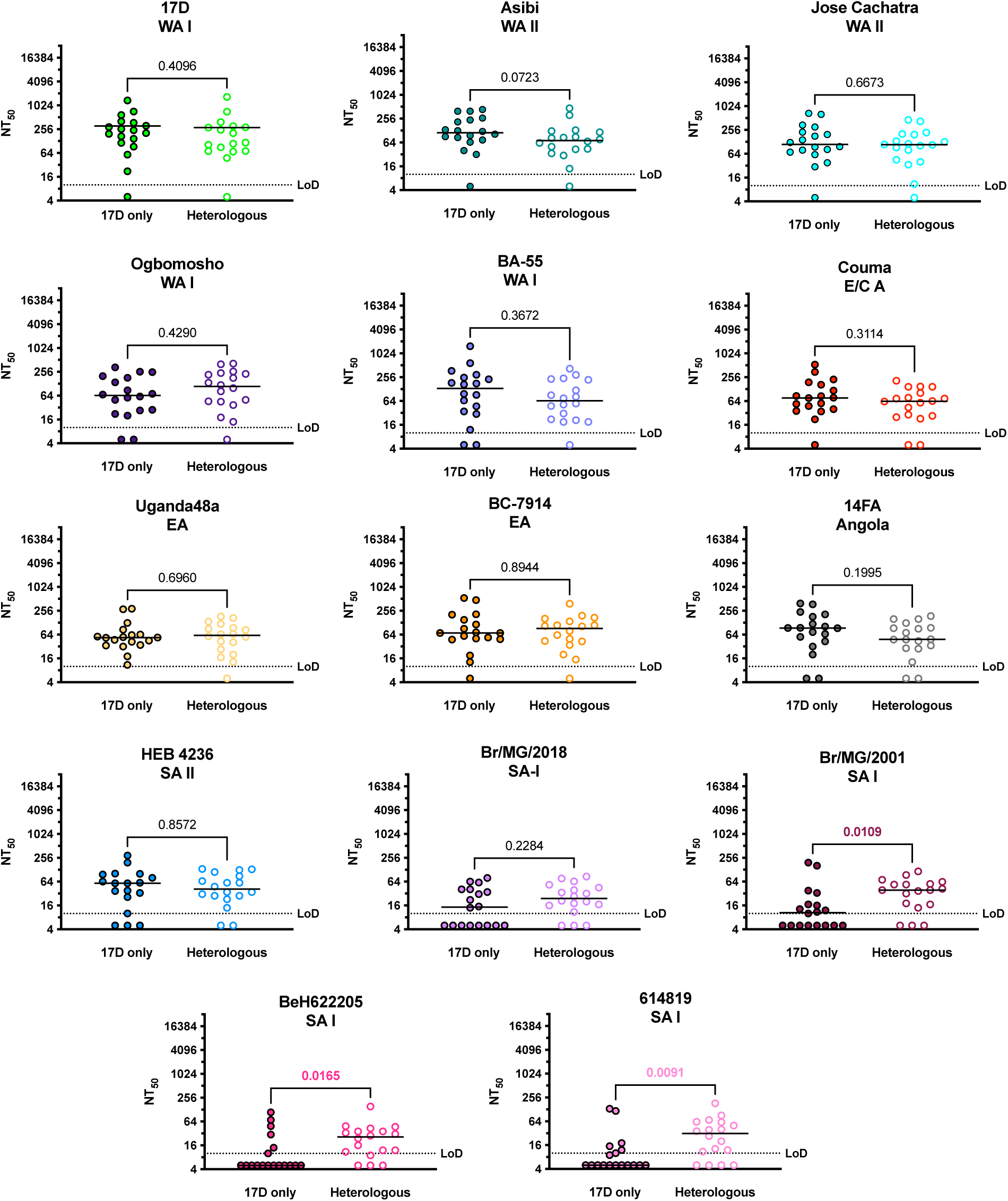
Heterologous vaccinees have increased potency against SA-I strains compared to 17D-only vaccinees. Individual plots show GMTs of 17D-only-(open circles, right) and heterologous vaccinees (filled circles, left) against a single virus, with virus strain and genotype indicated at the top. Lower limit of detection (dotted line) is 1:10 and upper limit of detection is 1:2560. Horizontal lines indicate medians. P-values derived from Mann-Whitney U; statistically significant p-values are in bold and color.

To further investigate the increased potency and breadth of NAbs observed amongst heterologous vaccinees, we stratified GMTs by *Orthoflavivirus* infection history and compared pooled GMTs (Figure 4A and C). Among 17D-only vaccinees, GMTs against Uganda48a (adj. p=0.0214), HEB4263 (adj. p=0.0006), and SA-I strains Br/MG/2001 (p<0.0001, BeH622205 (adj. p<0.0001), 614819 (adj. p<0.0001), and Br/MG/2018 (p adj. <0.0001) were significantly reduced compared to 17D. (Figure 4A). Amongst heterologous vaccinees, GMTs against Uganda48a (adj. p=0.0300), 14FA (adj. p=0.0200), Couma (adj. p=0.0086), HEB4236 (adj. p=0.0008), Br/MG/2001 (adj. p<0.0001), BeH622205 (adj. p<0.0001), 614819 (adj. p<0.0001, and Br/MG/2018 (adj. p<0.0001) were significantly reduced compared to 17D (Figure 4C).

**Figure 4:**
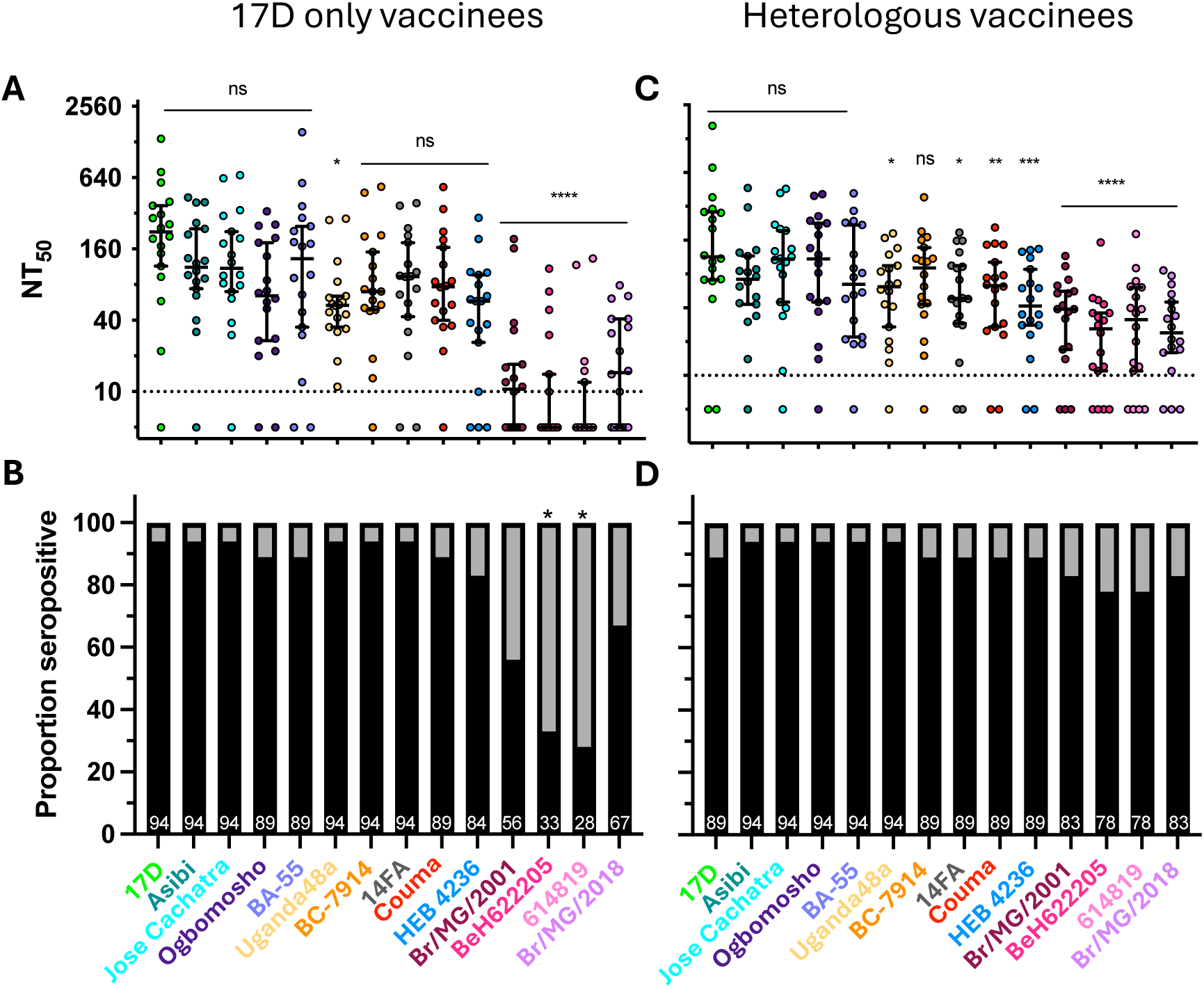
Heterologous vaccinees have increased seropositivity to SA-I strains. NT_50_ against each virus **(top panels)** for **(A)** 17D only vaccinees and **(C)** heterologous vaccinees. Each dot represents the geometric mean titer (GMT) of a single participant sera, calculated from biological duplicates. Horizontal bars and whiskers are geometric means with 95% confidence intervals. Lower limit of detection (dotted line) is 1:10 and upper limit of detection is 1:2560. GMTs <1:10 were given an arbitrary value of 5. Virus strains are color-coded according to the key in Figure 2 and arranged by genotype. P-values derived from Friedman test with Dunn’s multiple comparison: * adj. p<0.05, ** adj. p<0.01, *** adj. p<0.001, **** p<0.0001. Bar graphs showing proportion of seropositive (NT_50_ ≥1:10, black) and seronegative (NT_50_ <1:10, grey) **(bottom panels)** amongst **(B)** 17D only-and **(D)** heterologous vaccinees. White numbers indicate percentage of total (n=18). P-values derived from analysis of means of proportions (ANOMP): * indicates p<0.001.

Next, we calculated fold-differences between pooled GMTs for every virus pair (Figure 5). Overall, fewer significant differences were observed between pooled GMTs amongst heterologous vaccinees (20/91 pairs) (Figure 5B) compared to 17D-only vaccinees (34/91pairs) (Figure 5A). GMTs were significantly reduced for SA-I strains—Br/MG/2001, BeH622205, 614819, and Br/MG/2018—against most other viruses for 17D-only vaccinees with fold-differences ranging 1.0-20.7 and 32/46 significant differences between virus-pairs (Figure 5A). In contrast, fewer significant differences were observed for SA-I strains amongst heterologous vaccinees (18/46 pairs) ranging 1.0-6.0 (Figure 5B), supporting our previous observation that heterologous infection increases potency and breadth of 17D-immune sera.

**Figure 5:**
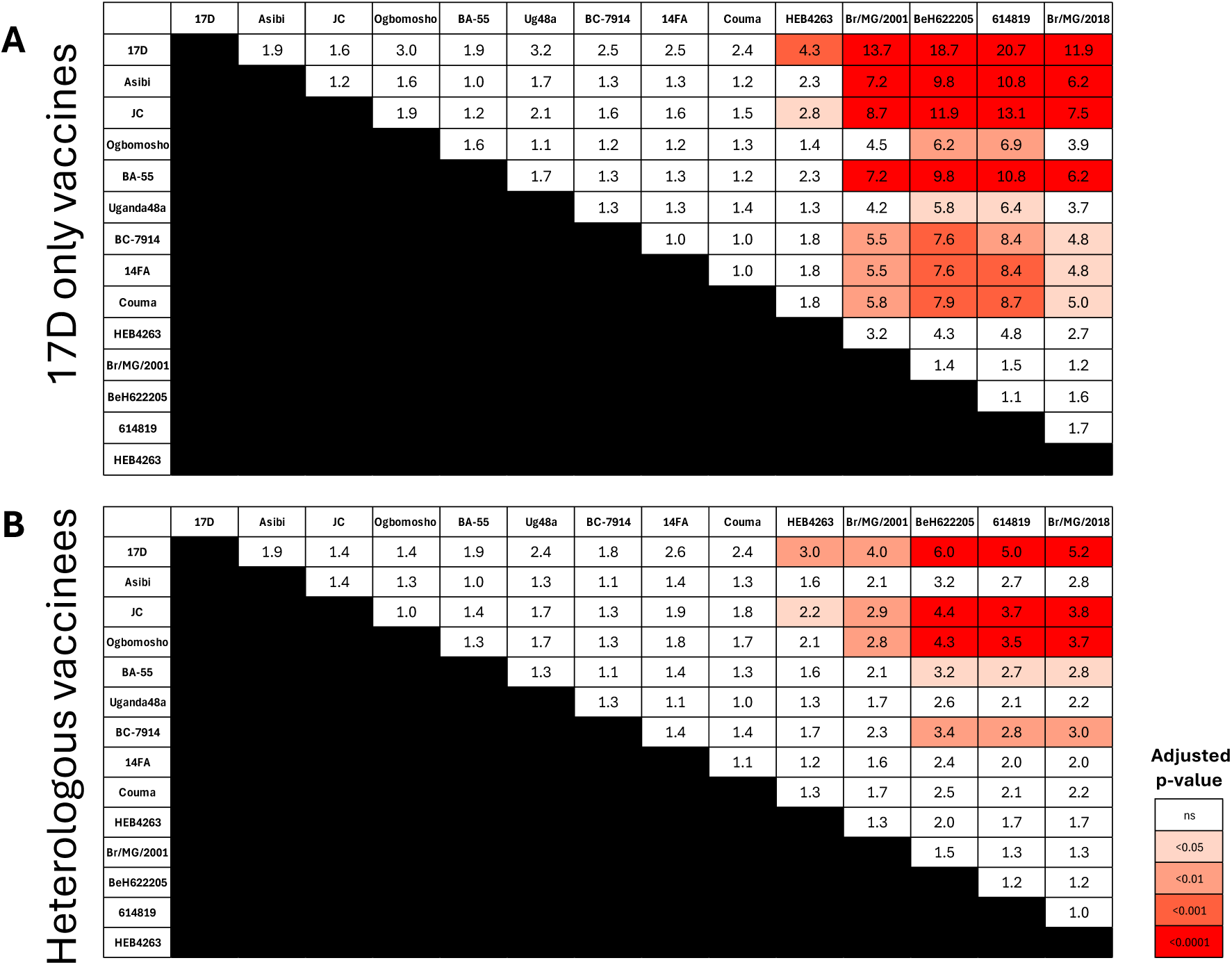
Heterologous have reduced GMT fold-differences between wild-viruses and 17D. Matrix showing fold differences between NT_50_ GMTs (horizontal bars, Figure 4A) between viruses, amongst **(A)** 17D-only vaccinees and **(B)** heterologous vaccinees. Boxes are colored by adj. p-values derived from Friedman test with Dunn’s multiple comparisons test.

### Vaccinees with heterologous flavivirus infection have increased seropositivity to SA-I strains

We next examined the impact of heterologous immunity on the proportion of vaccinees that were seropositive against each virus. By stratifying by *Orthoflavivirus* infection history, we observed significantly reduced proportions of seropositive vaccinees amongst 17D-only vaccinees, with 33% against BeH622205 (p<0.001), and 28% against 614819 (p<0.001) (Figure 4B). Both Br/MG/2001 (56%) and Br/MG/2018 (67%) also trended towards reduced proportion of seropositive vaccinees but were not significant (p>0.05). In contrast, heterologous vaccinee seropositivity against SA-I was not significantly reduced compared to other viruses, with 83% seropositive against Br/MG/2001, 78% against BeH622205, 78% against 614819, and 83% against Br/MG/2018 (Figure 4D), with ≥89% seropositive vaccinees for all other viruses. We re-examined FRNT_50_ values and Dayhoff distances by *Orthoflavivirus* infection history and found again strong negative correlations between WT virus amino acid distance from 17D and neutralization titers for both 17D-only vaccinees (Pearson R=-0.800, p=0.0005) and heterologous vaccinees (Pearson R=-0.810, p=0.0004).

We next evaluated the impact of vaccination-infection sequence. Seven participants were *Orthoflavivirus* infected before vaccination, 10 were infected after vaccination, and one was unknown, with no significant differences between these two groups regarding GMT (p>0.05) and seropositivity (p>0.05) (data not shown).

### SA-I strains form a distinct antigenic cluster

Antigenic cartography (AC) is a method used to visualize the antigenic relationship between virus strains using NT titers or similar tests of antigen recognition by immune sera, *i.e.* hemagglutination inhibition. AC was developed first for influenza^24^ and has subsequently been used to characterize DENV,^23, 26^ ZIKV^22^, and SARS CoV-2,^27^ among others. Antigenic maps are constructed by plotting virus strains and immune sera in a multi-dimensional map space, usually 2-D or 3-D. The coordinates of any given virus and serum on the map are the result of optimization that attempts to minimize the sum of squares between the table distances (derived from a matrix of neutralization titers) and map distances (straight-line distances) between any given virus-serum pair.^28^ The resulting map is a relational visualization of all viruses and all sera within the model where the proximity of objects suggests antigenic similarity and each box represents a two-fold serum dilution.^23^ Antigenic cartography offers a nuanced visualization of the antigenic relationship between viruses based on the functional readout of neutralization, which allows us to evaluate immune responses that may not be obvious from linear sequence data alone. The antigenic relationship of WT yellow fever viruses using antigenic cartography has not, to our knowledge, been previously explored. We constructed an antigenic map using GMTs against all 14 viruses for all sera (Figure 6A).

**Figure 6:**
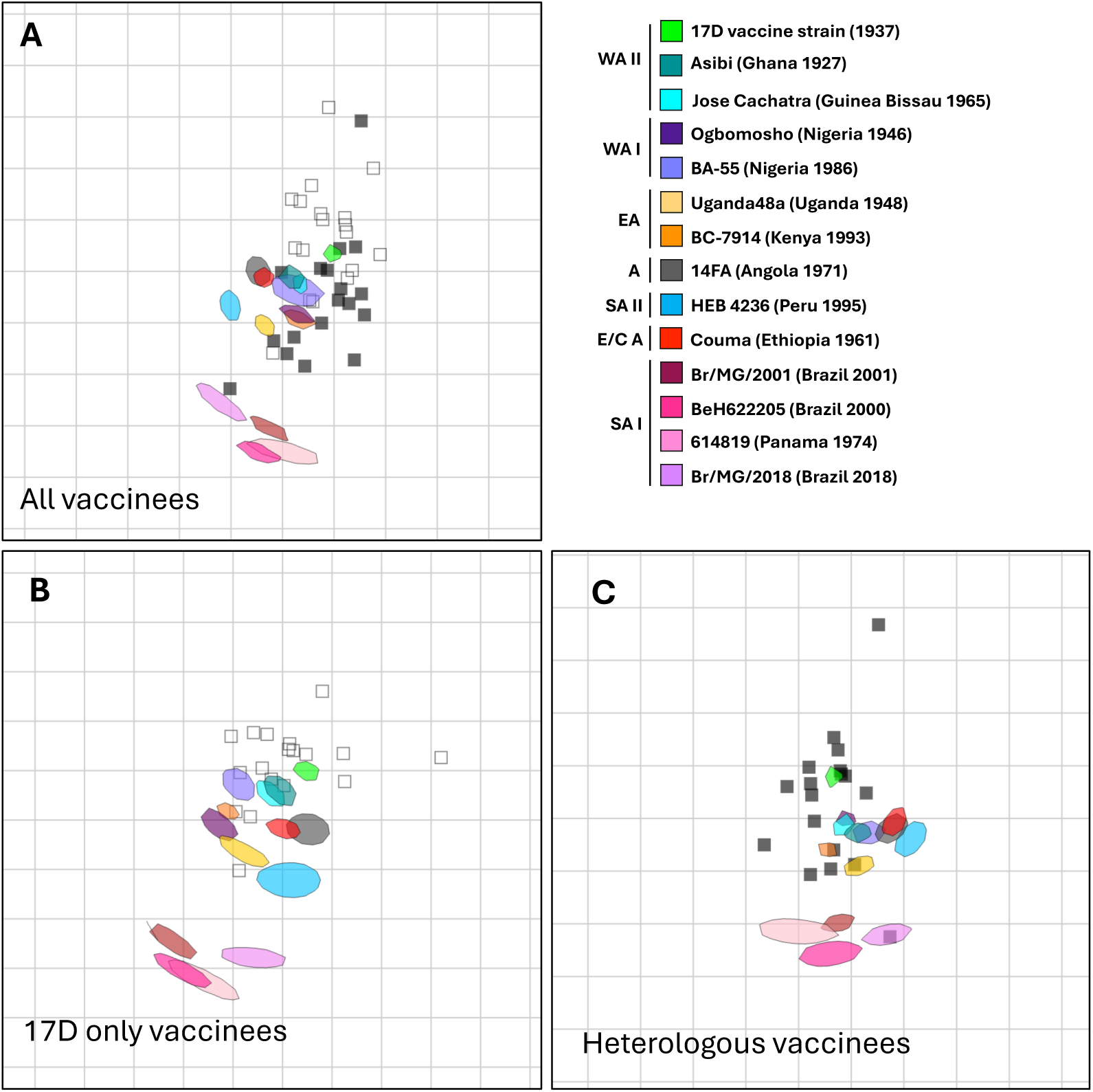
Antigenic cartography of 17D and wild-type YFV strains, using 17D-immune sera. Antigenic cartography maps generated using **(A)** all 17D-vaccinees sera (n=36), including 17D only-(open squares) and heterologous vaccinees (filled squares), **(B)** 17D only vaccinees, and **(C)** heterologous vaccinees. Maps were generated using the Racmacs package in R (version 4.3.1) and RStudio (version 2023.06.1+524). A single gridline-define box represents a two-fold serum dilution of a neutralization titer.

As expected, SA-I strain viruses were most antigenically distant from 17D and formed a distinct cluster. Antigenic map distances between virus strains and sera were greatest for SA-I strains at 3.16-3.69 antigenic units compared to 2.23 for HEB4263, and 1.00-1.75 for all other WT strains (Supplemental Table 2). To follow up our observation of differences inF RNT_50_ between 17D only and heterologous vaccinees we constructed antigenic maps using GMTs stratified by *Orthoflavivirus* infection history. The antigenic distances between sera and SA-I strains increased when constructed using 17D-only vaccinee GMTs (3.95-4.64) (Figure 6B, Supplemental Table 2) and decreased with heterologous vaccinee GMTs (2.26-2.88) (Figure 6C, Supplemental Table 2). Heterologous immune sera were also more centered within WT viruses compared to 17D-only vaccinees. Overall, these data suggest that heterologous immune sera recognize a wider breadth of antigenically diverse WT YFVs compared to 17D-only immune sera.

### Reduced potency against SA-I strains is observed at 28 days post-vaccination

To begin to investigate if the reduced potency 17D immune sera observed against the SA-I strains is the results of primary vaccine failure (failure to seroconvert following vaccination) or because of secondary vaccine failure (waning immunity following successful seroconversion) we performed FRNTs on 15 vaccinee sera at 28 days post-vaccination against a subset of our WT virus panel, including 17D, BA-55 (WA-I), and BeH622205 (SA-I) (Figure 7).

**Figure 7:**
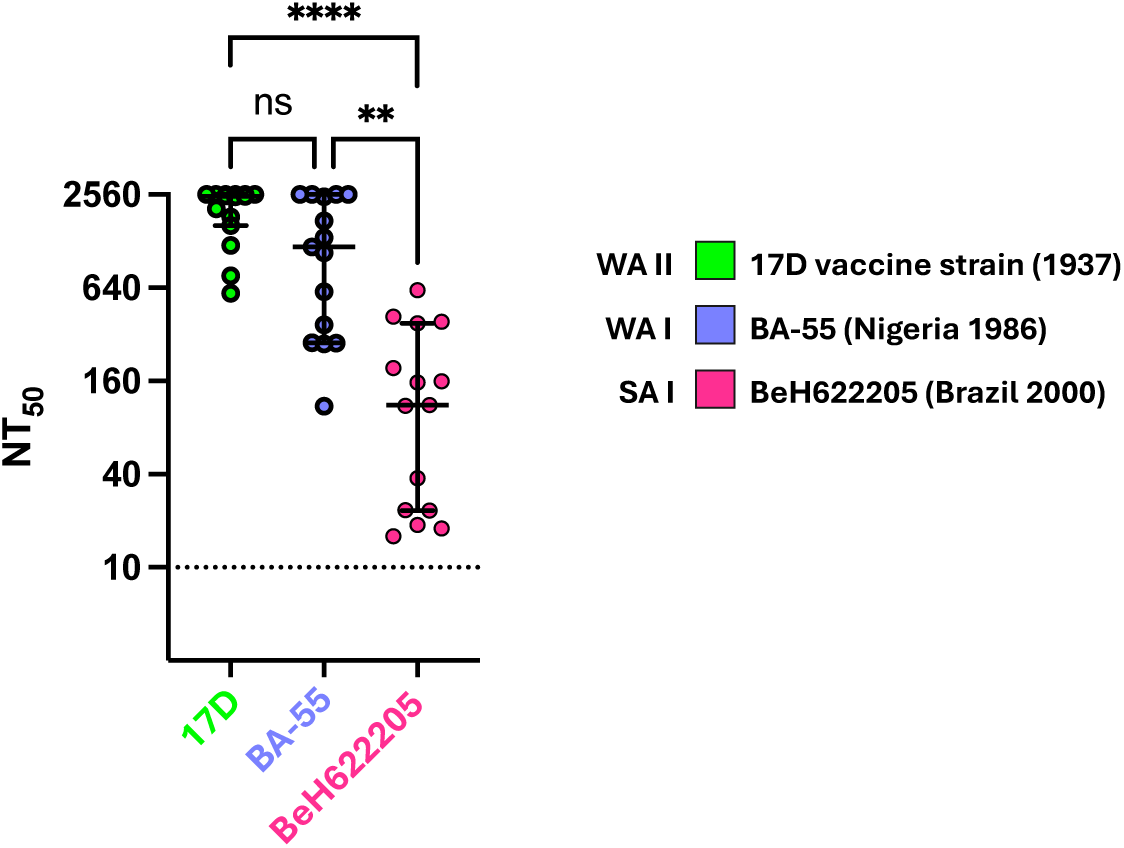
Reduced potency against the SA-I genotype is observed 28 days post-vaccination. NT_50_ GMTs of vaccinee sera 28 days post-vaccination against representative WT viruses. Each dot represents the geometric mean titer (GMT) of a single participant sera, calculated from biological duplicates. Lower limit of detection (dotted line) is 1:10 and upper limit of detection is 1:2560. GMTs <1:10 were given an arbitrary value of 5. P-values derived from Friedman test with Dunn’s multiple comparison: ** adj. p<0.01, and **** p<0.0001.

Study participants were aged 23-49 years (median 30) at vaccination and 43% female. Race/ethnicity data is omitted to maintain anonymity. As with our earlier data, we observed significantly reduced pooled GMTs against SA-I strain BeH622205 compared to 17D (adj. p<0.0001), and BA-55 (adj. p=0.0042) (Figure 7). All vaccinees were seropositive against all viruses, however, six participants had an FRNT_50_ of <1:40 against BeH622205 at 28 days post-vaccination. Based on this preliminary data, we predict secondary vaccine failure as the major mechanism contributing to the high proportion of seronegative vaccinees observed amongst our cross-sectional cohort (Figures 1 & 4), but do not rule out the possibility of primary vaccine failure amongst a larger cohort.

### YFV cross-reactivity of DENV immune sera from unvaccinated individuals

After finding increased potency and breadth of heterologous immune sera against WT YFV, we next assessed baseline cross-reactivity of DENV-immune sera, without 17D vaccination, against WT YFV, a comparison that has not, to our knowledge, been previously done. We tested 25 DENV-immune sera from individuals who denied prior 17D vaccination (Figure 8).

**Figure 8.**
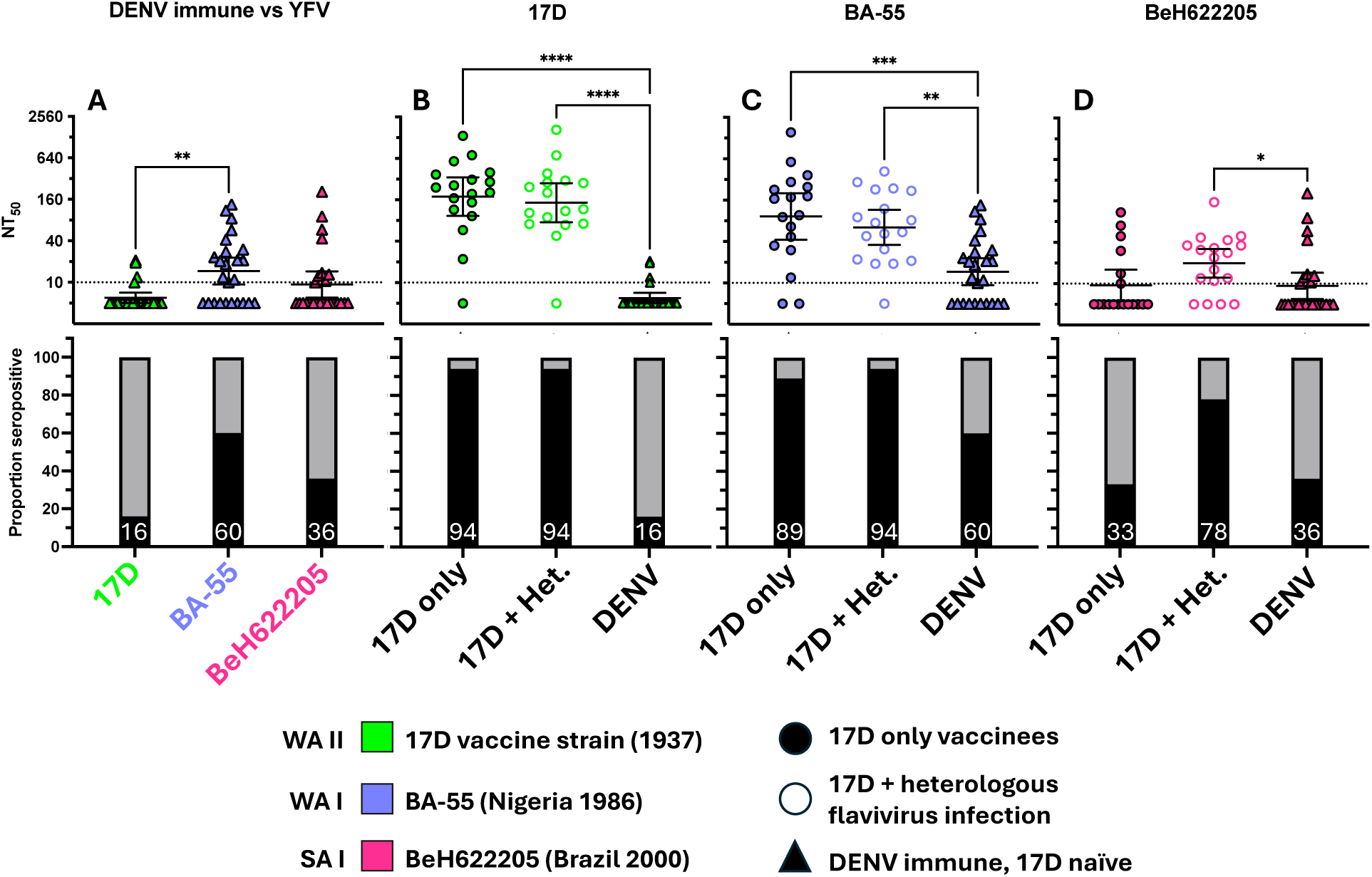
YFV cross-reactivity of DENV-immune unvaccinated individuals. NT_50_ against viruses **(top panels)**. Each dot represents the geometric mean titer (GMT) of a single participant sera, calculated from biological duplicates. Horizontal bars and whiskers are geometric means with 95% confidence intervals. Lower limit of detection (dotted line) is 1:10 and upper limit of detection is 1:2560. GMTs <1:10 were given an arbitrary value of 5. Virus strains are color-coded according to the key. P-values derived from Friedman test with Dunn’s multiple comparison: * adj. p<0.05, ** adj. p<0.01, *** adj. p<0.001, **** p<0.0001. Bar graphs **(bottom panels)** showing proportion of seropositive (NT_50_ ≥ 1:10, black) and seronegative (NT_50_ <1:10, grey) participants, with percentage of total (n=25) shown with white numbers. (A) DENV-immune sera against viruses, followed by 17D-only vaccinees (filled circles), heterologous vaccines (open circles), and DENV-immune sera (triangles) against **(B)**17D, **(C)** BA-55, and **(D)** BeH622050.

Four of 25 (16%) were seropositive against 17D, a significantly lower percentage than both 17D-only and heterologous immune vaccinees, with positive FRNT_50_ titers ranging from 1:12 to 1:21 (Figure 8A & B). Cross-reactivity was greatest against the WA-I strain BA-55, with 15 of 25 (60%) seropositive and a titer range from 1:10 to 1:135, but still significantly lower than both 17D-only vaccinees (adj. p=0.0001) and heterologous vaccinees (adj. p=0.0021) (Figure 8A & C). Finally, DENV-immune cross-reactivity against the SA-I strain BeH622205 was limited, with 9 of 25 (36%) seropositive with an FRNT_50_ range among the positives of 1:12 to 1:207 (Figure 3A & D). Interestingly, there was no significant difference between DENV-immune and 17D-only immune FRNT_50_ titers (adj. p=0.9999) but heterologous vaccinee GMTs were significantly higher than DENV-immune (adj. p=0.0220) (Figure 8D).

## Discussion

Historically, most studies establishing 17D-elicited NT titers have used the vaccine strain 17D and occasionally the parental Asibi strain.^9, 29, 30^ This constraint is logistical – WT YFV must be handled at a more resource intensive BSL-3 biocontainment level and assembling a test panel of viruses is non-trivial. 17D is a convenient, fully homotypic, test virus, and generalizability of 17D NT titers to NT titers against WT viruses has been long assumed but never broadly tested. Two recent studies have utilized an authentic SA-I strains isolated during the recent outbreak in Brazil or YFV reporter viral particles (RVPs),^17, 18^ to demonstrate decreased potency of 17D-vaccinee sera against individual SA-I strains: Haslwanter et al., 2022^17^ observed a ∼7-8-fold reduction between FRNT_50_ between 17DD and SA-I strain ES-504 (n=24) and Goncalves et al., 2024^18^ observed ∼2-3-fold reduction between 17DD and SA-I strain Hu-BR2018 (n=23) which are similar to the fold-reduction observed amongst out heterologous vaccinees (4.7-7.1) However, neither study examined other SA-I strains or other WT YFV genotypes.

Haslwanter et al., 2022^17^ also demonstrated decreased potency of 17D vaccinee sera amongst 16 non-endemic US vaccinees, but using RVPs, which may not fully recapitulate the antigenic landscape of authentic viruses. Using purified monoclonal antibodies and chimeric RPVs, the authors mapped two sites within domain II (EDII) and the DII-DI hinge domain, containing five amino acid substitutions between SA-I and African strains 17D and Asibi; H67N, A83E, D270E, N271S, and N272K, located within discontinuous sequence encoding EDII that appear responsible for the reduced neutralization observed against their SA-I strain by their immune sera panel. Using our expanded library of authentic viruses, including three SA-I strains, we confirmed all five amino acid substitutions in our SA-I strains: Br/MG/2001, BeH622205, BR/MG/2018, and 614819 which was isolated Panama in 1974, demonstrating that these mutations arose at least 42 years before the recent outbreak in Brazil. We also identified the N271S mutation in our representative SA-I strain HEB4263 which trended towards decreased GMTs and fewer seropositive vaccinees compared to 17D and other African WT viruses, supporting the previous finding that N271S alone sufficiently impedes neutralization by 17D-immune sera.^17^

To our knowledge, ours is the most extensive characterization of the potency and breadth of 17D-vaccinee sera with diverse vaccination and host immune backgrounds against the seven circulating WT YFV genotypes. We observed significantly reduced potency of vaccinee sera against WT YFVs compared to the 17D vaccine strain, with the most significant differences observed against SA-I strains, consistent with Hanslwanter^17^ and Goncalves.^18^

We also identified a key modulating host factor—serologic evidence of heterologous *Orthoflavivirus* infection—that significantly increased the potency and breath of 17D-immune sera against SA-I strains and the proportion of vaccinees who were seropositive against SA-I. These data suggest that heterologous *Orthoflavivirus* infection may effectively “boost” the immunity elicited by the 17D vaccine, a finding that has implications for 17D vaccination strategies between endemic and non-endemic populations. Interestingly, Haslwanter et al. found 88% (21/24) of Brazilian vaccinees to be seropositive against the SA-I strain ES-504, a rate similar to that observed amongst our heterologous vaccinees against SA-I strains (72-83%).

Whilst Haslwanter et al. did not characterize *Orthoflavivirus* infection history in their Brazilian cohort, DENV is endemic in Brazil and Brazil experienced an unprecedented ZIKV outbreak 2015-2016, making it possible that their cohort included heterologous vaccinees. Additionally, a recent report by Shinde et al.^31^ found that prior DENV or ZIKV immunity suppressed YFV viremia in a WT YFV challenge model in cynomolgus macaques. The authors concluded that DENV and ZIKV immunity may potentially limit transmission of YFV in urban settings where DENV and ZIKV seroprevalence is high.

Antigenic cartography is a powerful tool used for surveillance of circulating viruses and to guide, for example, influenza vaccine formulation,^24^ but has not been previously employed to characterize the antigenic landscape of WT YFV. Here we constructed the first antigenic map of WT YFVs, finding that SA-I strains form a distinct antigenic cluster and that heterologous vaccinee sera has reduced antigenic distance to SA-I strains compared to 17D-only vaccinees, suggesting the potential for increased 17D vaccine efficacy amongst heterologous vaccinees.

Beyond defining the antigenic landscape of YFV, continued application of antigenic cartography could enhance surveillance of circulating YFV strains and facilitate the prediction regarding risk of an outbreak within a specific population and potentially inform future vaccine design strategies.

Our work does not establish the basis for increased potency of heterologous immune sera against SA-I strains. However, we note that the N67 mutation in SA-I WT YFV is otherwise unique to and highly conserved across DENV serotypes. For DENV, N67 contributes to a glycosylation site that has been shown to interact with host cell receptor dendritic cell-specific ICAM3 grabbing nonintegrin (DC-SIGN),^32^ is required for DENV infectivity, and has been implicated in pathogenesis,^33^ although the N67 mutation in SA-1 does not introduce a new glycosylation site.

Ours is the first report of the cross-neutralization activity of DENV-immune sera against WT YFV. We found evidence of cross-neutralization of WT YFV by DENV-immune sera, with low-to mid-level cross-neutralization by some DENV-only immune sera against WT YFV. However, the extent and magnitude of the cross-reactivity alone does not explain the greater potency and breadth of heterologous vaccinee sera against SA-I YFV strains. It is striking that 26% of our DENV-only participant sera include antibodies that neutralized Br/MG/2001, and we hypothesize that DENV infection results in the generation of memory B cells capable of recognizing N67 epitopes that have low level cross-reactivity to YFV SA-I strains, even in the absence of 17D vaccination. Similarly, we suspect that vaccination with 17D elicits memory B cells with low level cross-reactivity against DENV strains, and that the combination of DENV infection and 17D vaccination provide secondary antigen recognition, affinity maturation resulting in differentiation of memory B cells into progeny plasma cells that secrete antibodies with increased cross-reactivity to N67 epitopes and therefore SA-I strains.

Strengths of our study include the use of a diverse, authentic WT YFV panel, enabling us to establish potency and breadth of the previously unexplored WT YFV antigenic landscape. Within our cohort, we found a balanced distribution of years post vaccination, and a distribution of ages at vaccination that is particularly representative of traveler populations worldwide.

Additionally, the balanced distribution of heterologous-and 17D-only vaccinees which was confirmed by serology provides confidence in our finding that heterologous infection plays a role in boosting 17D immunity.

Our study’s limitations include a relatively small cohort, lacking children, or individuals vaccinated over the age of 69 years, and overrepresented by white and female participants and some statistical analyses, particularly of our smaller heterologous and 17D-only vaccinee subsets, should be interpreted cautiously. We found significantly decreased seropositivity against SA-I strains suggesting an increased rate of vaccine failure against SA-I strains, however it is not known if this translates to increased risk of infection. Additionally, our antigenic maps were constructed exclusively using 17D vaccinee immune sera which deviates from typical antigenic cartography utilizing immune sera and homologous viruses, for example, to properly define SA-I strains, sera from individuals naturally infected with an SA-I virus should be included.

Importantly, we did not include vaccinees who were >11 years post vaccination in our study, and so our findings cannot be generalized beyond this interval.

Future studies to validate our findings should include larger and more diverse cohorts to ensure generalizability to a broader population. Our data suggests the potential for increased risk of breakthrough infection with SA-I strains and calls for further evaluation of this risk. 17D breakthrough infections are not systematically reported^34^ and the extent to which breakthrough infections have or have not contributed to the large number of cases in South America in the past decade is not known, despite the serious public health implications of such breakthroughs. It is also possible that baseline DENV immunity in highly endemic countries like Brazil confers heterologous immunity which increases resistance to SA-I outbreaks; however, this has not been explored. With recent outbreaks and ongoing sylvatic transmission that maintains viral reservoirs within which viral evolution continues, YFV continues to pose a threat to public and global health. Within this context, our study amongst many others’ raises the question: is it time to evaluate the need to update this historical and important vaccine?

## Data Availability

All data produced in the present study are available upon reasonable request to the authors.

## Acknowledgements

We would like to acknowledge Brian Booty and Peter Sullivan from OHSU, and Tara Beatty, Evelin Coto, Lantoria Davis, Stephen Fortmann, Tarika Holness, Kristin Muessig, Elizabeth Shuster, and Mica Werner from KPCHR for their contributions to study coordination for the cohorts in this study. We also acknowledge the World Reference Center for Emerging Viruses and Arboviruses for the provision of virus strains Couma, Uganda48a, Ogbomosho, Jose Cachatra, HEB4236, and Asibi. Additionally, we acknowledge the Center for Disease Control Arbovirus Reference Collection for the provision of virus strains Br/MG/2001, BeH622205, BC-7914, BA-55, 614819, and 14FA. Thank you to Betania Drumond, PhD, Universidade Federal de Minas Gerais, and to Alec Hirsch, PhD, OHSU, Vaccine and Gene Therapy Institute for generously providing the 321/Br/MG/2018 virus and the 4G2 antibody, respectively.

