## Supplemental Tables for "New insights into an old vaccine: potency and breadth of neutralizing antibodies elicited by the yellow fever vaccine 17D are boosted by heterologous *Orthoflavivirus* infection"

Supplemental Material

| **Primer** | **Sequence 5’-3’** |
| --- | --- |
| **857R_YFVx** | AGCCATCTCTCAATCTTYTGS |
| **996F_YFV** | YGACAGRGAYTTCATTGAGGG |
| **1300F_YFV** | GGGAAAGGRAGCATTGTGG |
| **1520R_YFV** | CAYTCCAGTGTRGCTTTYCC |
| **2144F_YFV** | ACAAAGARGGAAGYTCAATAGG |
| **2340R_YFV** | GACCTTTGTTATCCAGYTCAAGC |
| **2843F_YFV** | ATGGAAGCTTCATCATAGAYGG |
| **3010R_YFV** | CACCCARGATRGATCCATCR |
| **YFVAmp1A** | ACCTTCAAGAGGTGTTCAAGG |
| **YFVAmp2A** | CTCCTCCATTGTTCATCTCATG |
| **YFVSeq2A_R** | CTATCATCATGCTCACCAAMCC |
| **1805F_Couma** | GGTGGTCATGTCTCTT |
| **2466R_Couma** | GCACATCCTTGGTCTGCC |
| **3011R_Couma** | TGCACCTAGTATAGCC |

**Supplemental Table 1: YFV Sequencing primers**

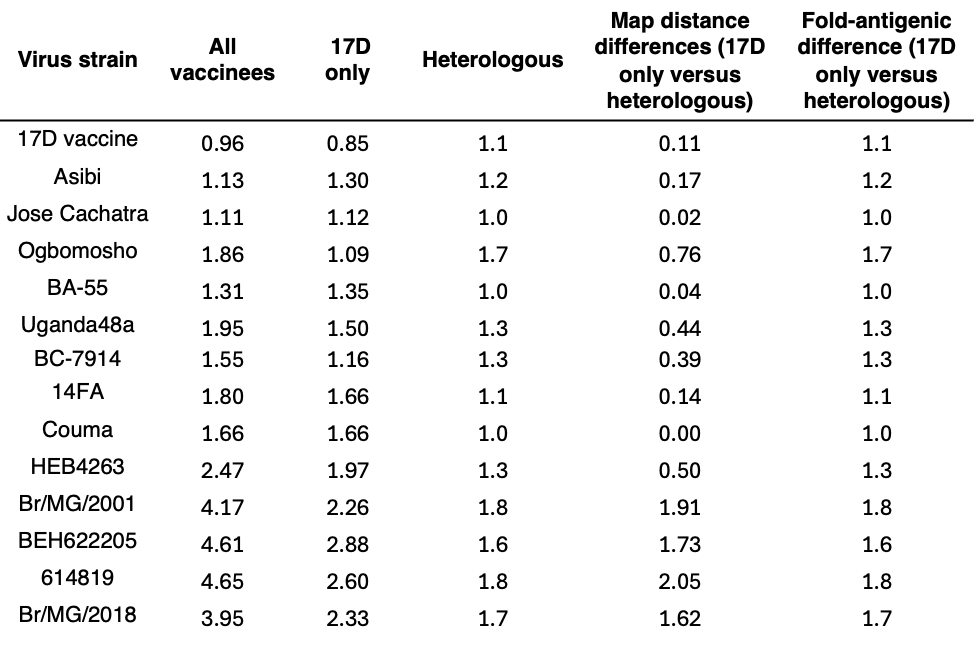

**Supplemental Table 2: Antigenic distances between virus strains and sera.** Table of median antigenic distances between sera and viruses, extracted from antigenic cartography models constructed using NT_50_ of total-, 17D-only-, and heterologous vaccinee sera (Figure 6A, B, and C). Values represent antigenic units corresponding to a single box within the antigenic map, where each box represents a two-fold serum dilution of a neutralization titer.
